# Brain morphology mediators of the link between childhood trauma and bipolar disorder: a large-scale international analysis

**DOI:** 10.1101/2025.11.02.25339336

**Authors:** Leonardo Tozzi, Maria R. Dauvermann, Emma Corley, Andrea Fernandes, Melody J. Y. Kang, Yanghee Im, Leila Nabulsi, Roberto Goya-Maldonado, Klaus Berger, Marco Hermesdorf, Hilary P. Blumberg, Lejla Colic, Tamsyn E. Van Rheenen, Elysha Ringin, Susan L. Rossell, James A. Karantonis, Lisa S. Furlong, Tilo Kircher, Frederike Stein, Udo Dannlowski, Dominik Grotegerd, Jair C. Soares, Mon-Ju Wu, Giovana B. Zunta-Soares, Benson Mwangi, Joaquim Radua, Enric Vilajosana, Eduard Vieta, Melissa J. Green, Emilie Olie, Guillaume Clain, Francesco Benedetti, Elisa Melloni, Beatrice Bravi, Delfina Janiri, Daniela Vecchio, Fabrizio Piras, Nerisa Banaj, Gabriele Sani, Gloria Roberts, Janice M. Fullerton, Bronwyn J. Overs, Philip B. Mitchell, Jonathan Savitz, Lakshmi N. Yatham, Josselin Houenou, Marion Leboyer, Amanda Rodrigue, David C. Glahn, Sophia I. Thomopoulos, Neda Jahanshad, Paul M. Thompson, Ole A. Andreassen, Christopher R. K. Ching, Dara M. Cannon, Yann Quidé, The ENIGMA Bipolar Disorder Working Group

## Abstract

Childhood trauma is a risk factor for bipolar disorder, but the biological mechanisms of this association remain incompletely defined. Gray matter differences observed after trauma exposure overlap with those reported in bipolar disorder, suggesting that the association between childhood trauma and bipolar disorder might be mediated through brain morphology.

Our goal was to determine whether cortical thickness, cortical surface or subcortical volume mediate the association between childhood trauma and bipolar disorder. We leveraged a large multi-site dataset from the ENIGMA Bipolar Disorder Working Group, comprising of 1,031 participants with bipolar disorder and 2,221 controls from 19 international cohorts. To identify brain morphology mediators of the association of childhood trauma and bipolar disorder, we used high-dimensional mediation analysis and validated our results using leave-one-site-out cross-validation and permutation testing for significance.

Severity of childhood trauma was directly associated with higher likelihood of having a bipolar disorder diagnosis (median coefficient 0.841, 95% CI: [0.834, 0.851], p<0.001). Significant mediators were hippocampal volume (0.004, 95% CI: [0.002, 0.005], p<0.001), medial orbitofrontal gray matter thickness (0.002, 95% CI: [0.002, 0.003], p<0.001), and superior frontal gyrus gray matter thickness (0.002, 95% CI: [0, 0.005], p<0.001).

Our results show that the severity of childhood trauma exposure is associated with bipolar disorder diagnosis in part through a smaller hippocampus, thinner cortex in the medial orbitofrontal gyrus and thinner cortex in the superior frontal gyrus. The identification of this mechanistic pathway improves our etiologic understanding of bipolar disorder and could help to identify those at risk and enable the development of new interventions.

## Introduction

Childhood trauma is associated with a higher likelihood of developing bipolar disorder, but the biological mechanisms involved in this association remain only partially understood ^1,2^. One hypothesis is that severe stress in childhood may induce lasting changes in the brain, which in turn may increase the likelihood of developing bipolar disorder later in life. This is supported by evidence that neurobiological changes related to childhood trauma overlap with brain alterations observed in adults with established bipolar disorder ^3^.

Adults with bipolar disorder show widespread thinner cortical gray matter, most pronounced in the prefrontal, middle frontal and fusiform regions ^4^. In addition to these cortical alterations, a bipolar disorder diagnosis is associated with smaller subcortical limbic structures such as the hippocampus and thalamus ^5^. Overall, the altered cortical and subcortical regions in bipolar disorder may be directly involved in the symptoms of the illness, including emotion regulation ^2^. Individuals exposed to childhood trauma show reduced gray matter in a pattern of partially overlapping regions. For example, large-scale neuroimaging studies and meta-analyses indicate that childhood trauma is associated with smaller gray matter volume in prefrontal and temporal areas, as well as amygdala, hippocampus and thalamus ^6–9^.

Several studies have investigated the impact of childhood trauma specifically in individuals with bipolar disorder and found that those exposed to childhood trauma show more severe clinical features and pronounced gray matter deficits compared to those not exposed to trauma in the pre-frontal, orbito-frontal, para-central cortices, hippocampus, thalamus and amygdala ^3,6,10–17^. These studies suggest that childhood trauma might increase the risk of developing bipolar disorder by reducing gray matter in these specific brain structures. However, most of these studies did not formally investigate a directed model that explicitly tested whether the early adversity may lead to increased risk of developing bipolar disorder through gray matter loss.

Here, we leverage a large multi-site dataset of 1,031 participants with bipolar disorder and 2,221 controls from 19 international cohorts to conduct the largest study to date investigating the relationship between the severity of childhood trauma exposure, brain morphology and bipolar disorder. We address the limitations of previous modeling strategies by building a mediation model in which childhood trauma’s association with bipolar disorder occurs via its effect on brain structure. This allows us to move from identifying simple associations to identifying potential directed mechanistic pathways leading from childhood trauma to bipolar disorder. Importantly, we take advantage of the multiple-site design to rigorously validate our results using leave-one-site-out cross-validation.

Based on previous findings, we expected the severity of childhood trauma exposure to be robustly associated with both thinner cortical gray matter and smaller subcortical volume of regions involved in emotion regulation. We expected these reductions to be associated, in turn, with an increased likelihood of having a bipolar disorder diagnosis.

## Methods

### Dataset

We analyzed data from the Enhancing NeuroImaging Genetics through Meta-Analyses (ENIGMA)-Bipolar Disorder Working Group (ENIGMA-BD), the largest international consortium studying neuroimaging signatures of bipolar disorder ^18^. ENIGMA-BD applies standardized neuroimaging, quality control and analysis pipelines to independently collect and process structural MRI data from around the world (https://github.com/ENIGMA-git/ENIGMA-FreeSurfer-protocol, https://enigma.ini.usc.edu/protocols/imaging-protocols). The dataset consisted of 2,224 healthy participants and 1,034 participants with bipolar disorder collected from 19 studies (Table 1). Three of the studies were conducted at the same site (Münster, Germany) but to account for differences in sample composition, study design and year of acquisition, we treated these data as if they were coming from separate sites for our leave-one-site-out cross validation analysis. For enrollment criteria and MRI sequences at each site, see Supplementary Tables S1 and S2. For a map of the participating sites’ locations, see Supplementary Figure 1.

**Table 1:**
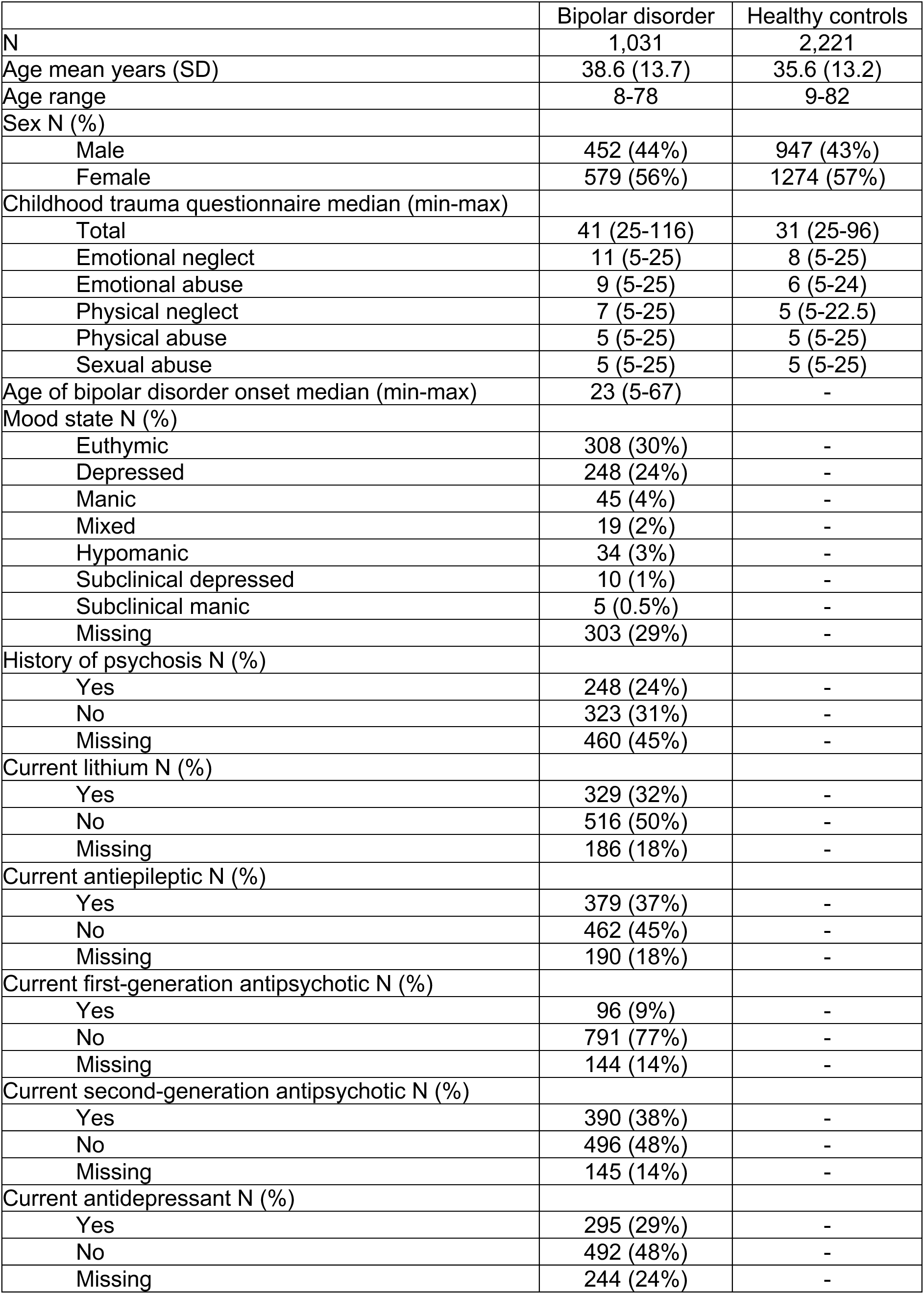
Demographics and clinical information of the dataset.

|  | Bipolar disorder | Healthy controls |
| --- | --- | --- |
| N | 1,031 | 2,221 |
| Age mean years (SD) | 38.6 (13.7) | 35.6 (13.2) |
| Age range | 8-78 | 9-82 |
| Sex N (%) |  |  |
| Male | 452 (44%) | 947 (43%) |
| Female | 579 (56%) | 1274 (57%) |
| Childhood trauma questionnaire median (min-max) |  |  |
| Total | 41 (25-116) | 31 (25-96) |
| Emotional neglect | 11 (5-25) | 8 (5-25) |
| Emotional abuse | 9 (5-25) | 6 (5-24) |
| Physical neglect | 7 (5-25) | 5 (5-22.5) |
| Physical abuse | 5 (5-25) | 5 (5-25) |
| Sexual abuse | 5 (5-25) | 5 (5-25) |
| Age of bipolar disorder onset median (min-max) | 23 (5-67) | - |
| Mood state N (%) |  |  |
| Euthymic | 308 (30%) | - |
| Depressed | 248 (24%) | - |
| Manic | 45 (4%) | - |
| Mixed | 19 (2%) | - |
| Hypomanic | 34 (3%) | - |
| Subclinical depressed | 10 (1%) | - |
| Subclinical manic | 5 (0.5%) | - |
| Missing | 303 (29%) | - |
| History of psychosis N (%) |  |  |
| Yes | 248 (24%) | - |
| No | 323 (31%) | - |
| Missing | 460 (45%) | - |
| Current lithium N (%) |  |  |
| Yes | 329 (32%) | - |
| No | 516 (50%) | - |
| Missing | 186 (18%) | - |
| Current antiepileptic N (%) |  |  |
| Yes | 379 (37%) | - |
| No | 462 (45%) | - |
| Missing | 190 (18%) | - |
| Current first-generation antipsychotic N (%) |  |  |
| Yes | 96 (9%) | - |
| No | 791 (77%) | - |
| Missing | 144 (14%) | - |
| Current second-generation antipsychotic N (%) |  |  |
| Yes | 390 (38%) | - |
| No | 496 (48%) | - |
| Missing | 145 (14%) | - |
| Current antidepressant N (%) |  |  |
| Yes | 295 (29%) | - |
| No | 492 (48%) | - |
| Missing | 244 (24%) | - |

In this study, we only included sites that had measured the severity of childhood trauma exposure using the short-form Childhood Trauma Questionnaire (CTQ) ^19^. One of the sites used an attenuated version of the short-form CTQ with only 2 items per domain. Scores from this site were re-scaled to the minimum and maximum values of the standard short-form CTQ. The CTQ measures five types of childhood trauma: emotional neglect, emotional abuse, physical neglect, physical abuse and sexual abuse. For our main analysis we used the total CTQ score, but we also investigated each type of trauma separately.

The imaging variables used in our analyses were based on our prior work ^4,5,18,20^. They included cortical thickness and surface area of the 68 cortical regions of the Desikan-Killiany atlas ^21^ and volume of 14 subcortical regions (automated segmentation, *aseg*), both derived using FreeSurfer ^22^. All imaging data underwent standardized and open source visual quality control following ENIGMA’s established protocols (https://github.com/ENIGMA-git, https://enigma.ini.usc.edu/protocols/imaging-protocols). All participating research sites obtained approval from local ethics committees, and all participants gave informed written consent. All procedures contributing to this work complied with the ethical standards of the relevant national and institutional committees on human experimentation and with the Helsinki Declaration of 1975, as revised in 2008.

### Imputation of missing values

Six participants had missing imaging data and were excluded, bringing the final dataset to 2,221 healthy participants and 1,031 participants with bipolar disorder. As a result of missing questionnaires and exclusion of gray matter measures after quality control, some variables were not available for a few participants: missingness was approximately 2% for CTQ and less than 10% for all imaging variables except measures from the right entorhinal cortex, left banks of the superior temporal sulcus and left superior temporal lobe (all 12%) (Supplementary Table 3). To maximize power and sample size, we imputed all missing values separately for each site by using multiple imputation by chained equations with random forests, using five iterations of a predictive mean matching model, as implemented in the R package miceRanger version 1.5.0 (10.32614/CRAN.package.miceRanger). The variables entered in this process were: age, sex, diagnosis, CTQ and non-missing gray matter measures.

We did not hypothesize that our effects of interest would be lateralized. Therefore, we calculated the mean of the right and left gray matter thickness and surface area (for cortical measures) or volume (for subcortical measures) for each brain region to obtain 75 bilateral-averaged measures, which we used in subsequent analyses.

### High-dimensional mediation analysis

#### Model specification

To investigate whether structural brain changes mediate the association between the severity of childhood trauma exposure and bipolar disorder, we employed high-dimensional mediation analysis (HIMA) ^23,24^.

HIMA implements a penalized regression framework that selects key mediators before estimating their indirect effects on the outcome. Specifically, a minimax concave penalty is applied to the outcome model to perform feature selection among the potential mediators, reducing the number of variables to those that contribute meaningfully to the relationship between the severity of childhood trauma exposure and bipolar disorder. The mediator models are then fitted using ordinary least squares regression for the remaining mediators.

We conducted HIMA using the *mediate_hima* function from the R package *hdmed* version 1.0.1 (10.32614/CRAN.package.hdmed). In our HIMA models, the predictor was the severity of childhood trauma exposure (CTQ total score), the mediators were the gray matter values from the 75 brain regions (averaged across left and right), and the binary outcome was diagnosis (bipolar disorder vs. healthy controls). We considered this temporal order of exposure (childhood trauma), mediators (brain morphology), and outcome (bipolar diagnosis) justified on developmental grounds ^25^. To account for differences in head size, we adjusted for the effect of intracranial volume on the volume of subcortical structures and cortical surface using linear regression separately for each site. We also included age and sex as covariates in the mediation models to account for their potential effects on brain structure and likelihood of bipolar disorder. Thus, we controlled for mediator-outcome and exposure-mediator confounds ^25^.

Previous studies found that certain types of childhood trauma might have specific effects on the likelihood of bipolar disorder ^2^. Therefore, we repeated our analysis separately using each of the five CTQ subscales as predictor (emotional neglect, emotional abuse, physical neglect, physical abuse and sexual abuse). To verify that our analyses were not driven by our missing value imputation procedure, we repeated our main analysis excluding records containing any missing values. Finally, to verify that the effects were not driven by children in the sample, we repeated our analysis on adults only (age ≥ 18 years).

#### Leave-one-site-out cross-validation

Given the multi-site nature of the ENIGMA dataset, we implemented a leave-one-site-out cross-validation approach to assess the generalizability of mediation effects across different research sites. For each site, the mediation model was trained using data from all other sites combined, while the held-out site served as the test set. This leave-one-site-out cross-validation framework ensured that the findings were not driven by site-specific biases.

For each cross-validation fold, first we applied ComBat correction within the training set to account for systematic differences between imaging data collected across different scanners and protocols. ComBat is an empirical Bayes method that adjusts for site-related batch effects while preserving biologically relevant variance ^26^. Here, the site effect correction was implemented using the combat.enigma package in R version 1.1.1 (doi: 10.32614/CRAN.package.combat.enigma), specifying diagnosis, childhood trauma, age and sex as effects to be preserved ^27^. Following this correction, all measures in the training set were standardized using a z-score transformation based on their mean and standard deviation. After these steps, a HIMA model was fit to the training set. Then, the resulting HIMA model was evaluated on the held-out test set. To do this, first the test set was standardized using the same mean and standard deviation used to standardize the training set. Then, the coefficients of the HIMA model fit on the training set were applied to the test set to obtain a predicted probability of bipolar disorder.

#### Model evaluation

Before interpreting the mediation model results, we evaluated the model’s ability to accurately classify bipolar disorder and healthy control individuals across the independent test sets. We considered an adequate predictive performance across independent test sets evidence that the model fitted the data well and was robust to site-specific variations. This gave us confidence to interpret mediation pathways and model coefficients. We quantified predictive performance on the test set using the area under the receiver operating characteristic curve. As a benchmark, we compared it to the published leave-one-site-out area under the curve of a model predicting bipolar disorder from ENIGMA imaging data alone in a sample of 853 individuals with bipolar disorder and 2,167 controls across 13 sites ^20^.

To assess significance of HIMA model coefficients, we used permutation testing. First, the observations for the mediators were randomly shuffled to break the relationship between predictor and mediators as well as between mediators and outcome. Then, the median of the mediator effects across sites was extracted. This procedure was conducted 1,000 times to build a null distribution of the mediator effects. We used the median across sites instead of the mean because the data was expected to be zero-inflated, since mediators can be set to zero by the HIMA regularization procedure. Then we compared the effects from the original mediation analysis to this null distribution to get a two-sided p-value for each mediator, defined as the fraction of null effects with an absolute value greater or equal from that of the null distribution. We only considered significant mediators for which p<0.05. The same procedure was used to assess significance of direct effects, by shuffling the childhood trauma, age and sex variables.

### Code availability

All code used in the analyses is publicly available on GitHub at https://github.com/leotozzi88/enigma_bd_mediation.

## Results

We conducted the HIMA in a large dataset of 1,031 participants with bipolar disorder and 2,221 controls (Table 1). Figure 1, Figure 2 and Table 2 show significant model effects. Supplementary Table 4 shows all model effects retained in at least one leave-one-site-out cross-validation fold.

**Figure 1:**
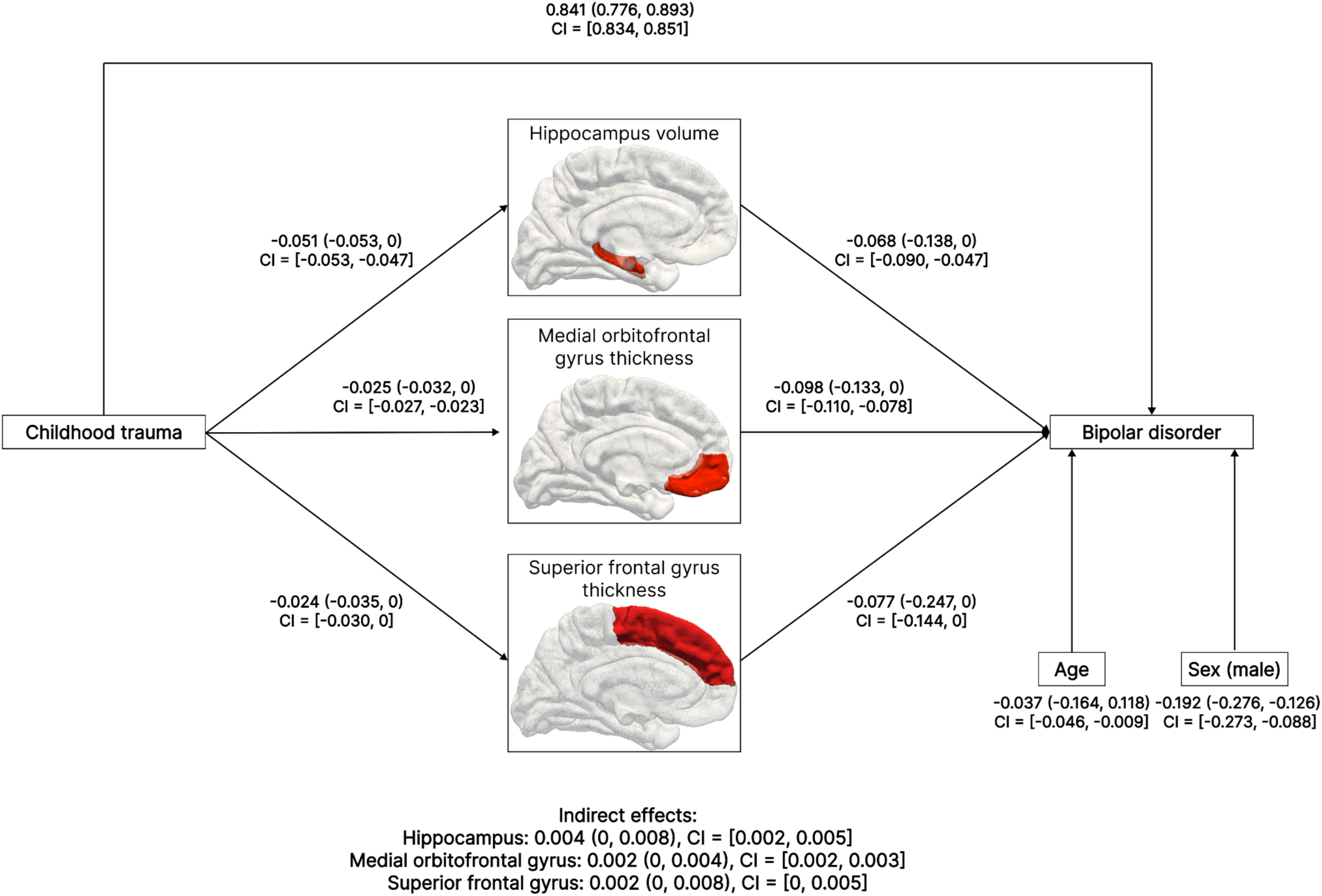
High-dimensional mediation analysis significant results. For each significant direct and indirect pathway, we show the median, minimum, maximum and 95% confidence intervals (CI) of the median across 19 leave-one-site-out cross-validation folds. We chose the median rather than the mean to account for the zero-inflated values generated by regularization. To determine significance, we generated a null distribution of the effects by permutation testing, used it to calculate a two-sided p-value and considered significant effects with p<0.05.

**Figure 2:**
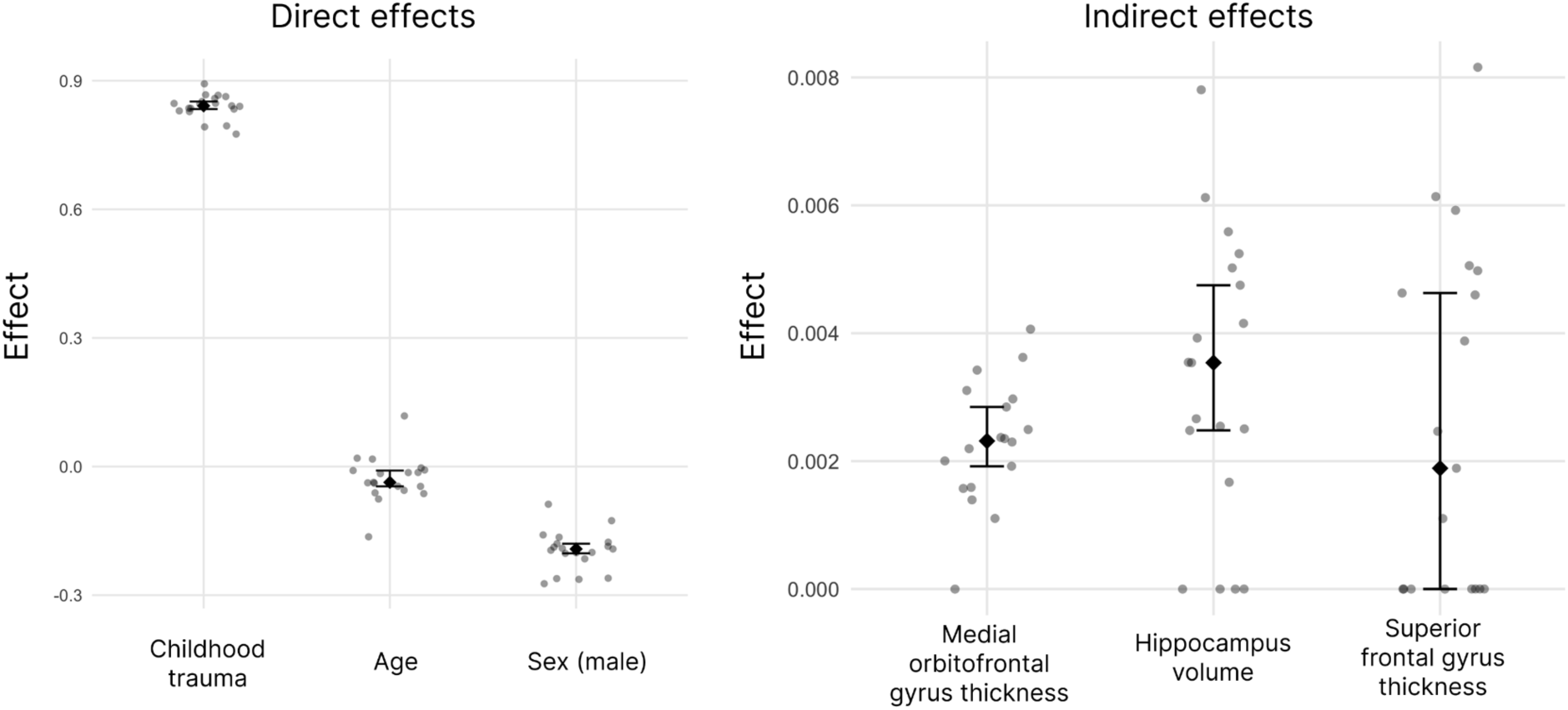
Direct and indirect effects in leave-one-site-out cross-validation folds. In 19 leave-one-site-out cross validation folds, we fit a high dimensional mediation model on data from all sites except one. We show the value of each significant direct and indirect effect in each fold (points), along with the median (diamonds) and CI of the median across folds (whiskers). We chose the median rather than the mean to account for the zero-inflated values generated by regularization. To determine significance, we generated a null distribution of the effects by permutation testing, used it to calculate a two-sided p-value and considered significant effects with p<0.05.

**Table 2:**
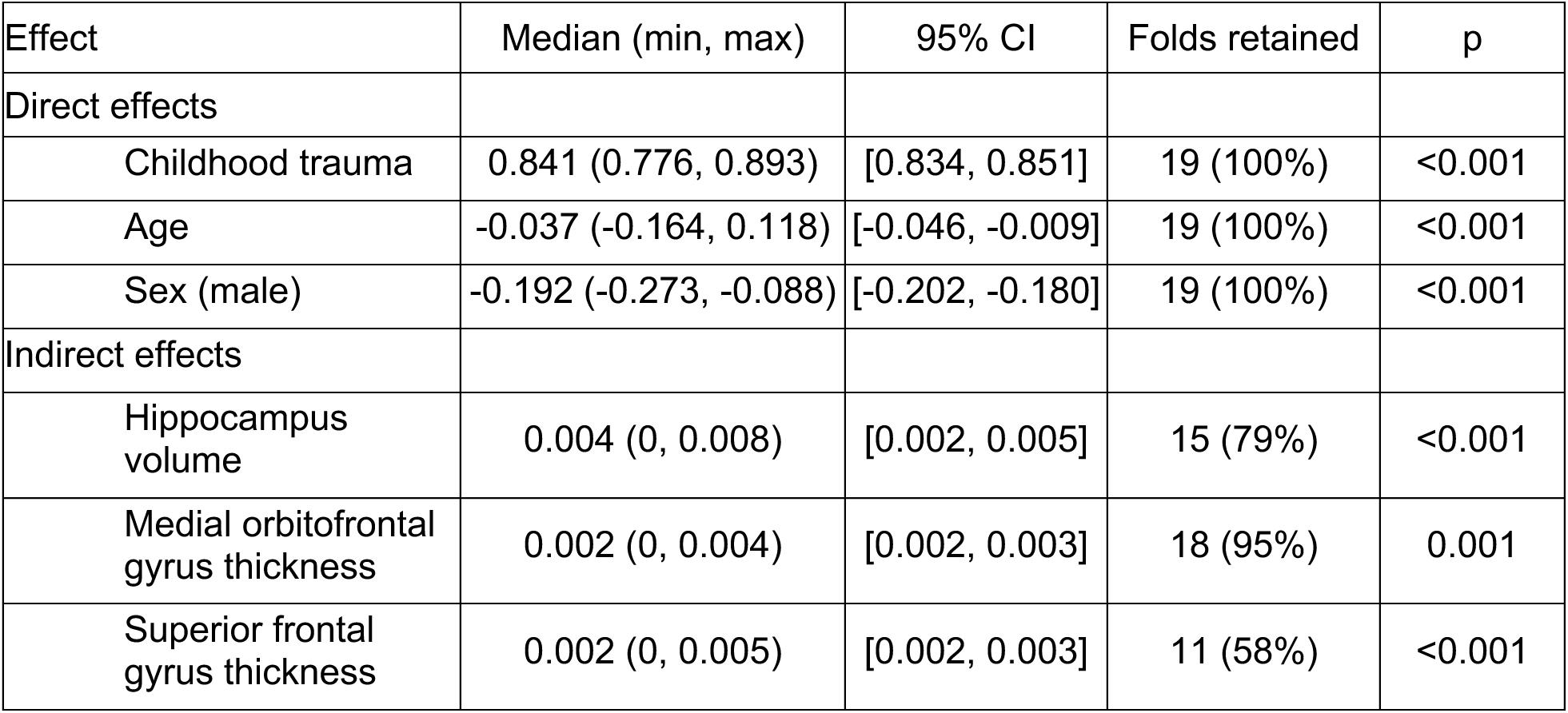
Direct and indirect high-dimensional mediation model effects. We show the median, minimum, maximum and 95% confidence interval (CI) of the median of each significant effect across 19 leave-one-site-out cross-validation folds. We chose the median rather than the mean to account for the zero-inflated values generated by regularization. We also show in how many folds the effect was retained in the feature selection procedure. To determine significance, we generated a null distribution of the effects by permutation testing, used it to calculate a two-sided p-value and considered significant effects with p<0.05.

### Model performance in predicting bipolar disorder

The HIMA model including CTQ as a predictor and brain morphology measures as mediators had good performance in predicting bipolar disorder on unseen data across leave-one-site-out cross-validation sites. It showed a mean area under the curve of 0.71 (CI: 0.67-0.76), with a higher non-overlapping confidence interval than the mean area under the curve of a previous model using only ENIGMA imaging data in a sample of 853 individuals with bipolar disorder and 2,167 controls across 13 sites (mean: 0.61, CI: 0.58–0.64) 20. Supplementary Figure 2 shows model performance at each site.

### Direct effects of childhood trauma, age and sex on bipolar disorder

All direct effects included in our analysis were significant. A higher CTQ total score increased the likelihood of a participant having a bipolar disorder diagnosis (median coefficient: 0.841, CI: [0.776, 0.893], folds retained: 100%). Males had a lower likelihood of having bipolar disorder compared to females (median coefficient: -0.192, CI: [-0.273, -0.088], folds retained: 100%). Older age was also associated with a slightly increased likelihood of having bipolar disorder (median coefficient: -0.037, CI: [-0.046, -0.009], folds retained: 100%).

### Brain mediators of childhood trauma on bipolar disorder

Three brain measures significantly partially mediated the relationship between childhood trauma and likelihood of a participant having a bipolar disorder diagnosis. Hippocampal volume showed the strongest effect on increased likelihood of bipolar disorder (median coefficient: 0.004, CI: [0.002, 0.005], folds retained: 79%), followed by thickness of the medial orbitofrontal gyrus (median coefficient: 0.002, CI: [0, 0.004], folds retained: 95%) and thickness of the superior frontal gyrus (median coefficient: 0.002, CI: [0, 0.005], folds retained: 58%).

Thickness of the fusiform gyrus, inferior frontal gyrus *pars opercularis*, inferior temporal gyrus, lingual gyrus and caudal middle frontal gyrus, surface area of the lingual gyrus and supramarginal gyrus and caudate volume were also retained as mediators during the feature selection process in some folds; however, these effects were not significant (Supplementary Table 4).

Types of childhood trauma showed weak to moderate correlation between each other (Spearman rho=0.28-0.57, Supplementary Table 5). When repeating the analysis on each type of childhood trauma separately, hippocampal volume remained a significant mediator of the relationships between every trauma type and likelihood of having a bipolar disorder diagnosis. For emotional neglect, emotional abuse and physical neglect, fusiform gyrus thickness was an additional mediator. For physical abuse and sexual abuse, inferior frontal gyrus *pars opercularis* thickness was retained as a mediator. Supplementary Tables 6-10 show model coefficients for the models including each type of childhood trauma separately.

To assess the impact of imputation, we repeated the analysis excluding records containing any missing values. Significant mediators did not change (Supplementary Table 11).

Finally, to assess the impact of including children, we repeated the analysis excluding participants < 18 years of age. Significant mediators did not change (Supplementary Table 12).

## Discussion

We conducted the largest neuroimaging analysis to date of the association between childhood trauma and bipolar disorder in a dataset containing 1,031 participants and 2,221 healthy controls. Our high-dimensional mediation analysis revealed that smaller hippocampal volume, thinner medial orbitofrontal cortex and thinner superior frontal gyrus cortex are potential pathways through which early-life trauma may increase risk for bipolar disorder.

Hippocampal volume emerged as the strongest significant partial mediator. This aligns with several previous findings that have shown smaller hippocampal volumes in individuals exposed to childhood trauma, both in clinical samples ^28–33^ and in the general population ^34,35^. Our prior ENIGMA-BD studies have found that individuals with bipolar disorder show smaller hippocampal volumes and hippocampal subfields, when childhood trauma was not accounted for ^5,36^. Here, we show that the impact of childhood trauma on hippocampal volume may be a key factor increasing risk of bipolar disorder. Future studies focusing on the hippocampus should therefore take into consideration the impact that childhood trauma exposure may have when interpreting differences in hippocampal morphology.

Thickness of the medial orbitofrontal gyrus and of the superior frontal gyrus were also significant mediators, in line with prior findings showing an association between reduced gray matter in the prefrontal cortex and childhood trauma ^37–41^. Thus, our findings further support the theory that childhood trauma is related to structural alterations in frontal brain regions, which are in turn associated with bipolar disorder. These regions are involved in emotion regulation and have shown differences in response to emotion-related tasks in individuals with bipolar disorder in functional neuroimaging studies ^42–45^

The hippocampus and prefrontal cortex, together with the amygdala, are the brain regions with the highest expression of high-affinity mineralocorticoid receptors, which are key in mediating the effects of stress hormones cortisol and corticosterone on the brain ^46,47^. It has been hypothesized that cumulative chronic stress due to childhood trauma during brain development may cause long-lasting changes in stress hormone axis regulation, which in turn may impact the brain and increase the risk of developing psychiatric disorders ^48,49^. Furthermore, the interaction between stress/trauma and genetic and/or familial risk of bipolar disorder may exacerbate the impact of trauma exposure in at-risk youth, leading to poor mental health outcomes ^50,51^. Our finding that the association between childhood trauma and bipolar disorder is mediated by the structure of the brain regions with the highest expression of high-affinity stress hormone receptors fits well within this view ^46,47^.

In our estimates of effect size, less than 1% of the association of childhood trauma and the likelihood of a bipolar disorder diagnosis was mediated by brain morphology. The small size of this effect could be due to several reasons. For example, our sample was not collected with the specific goal of investigating the relationship between childhood trauma and bipolar disorder, and thus individuals who had experienced intense high levels of childhood trauma were not specifically selected. Also, HIMA is an exploratory method involving regularization and it could have shrunk real effects to zero in some cross-validation folds. Even though the effects we found were small, we believe our findings are important, since they uncovered a biological pathway linking childhood trauma and bipolar disorder through three specific brain regions. Large scale collaborative efforts, such as ENIGMA, are key in enabling exploratory discovery of novel effects such as these. Future studies can build on our findings to investigate the pathway we identified with greater precision, for example by using approaches that measure type, severity, timing and repeat exposure of childhood trauma (e.g. using the Maltreatment and Abuse Chronology of Exposure or equivalent tools ^52^), collecting longitudinal clinical and multi-modal neuroimaging data, incorporating moderating or mediating effects on cognitive function, or gathering biomarkers of stress hormone axis and immune function.

It is noteworthy that the mediators we identified were similar across different trauma subtypes, with the exception of a significant mediation effect in the fusiform gyrus for emotional neglect, emotional abuse and physical neglect and a significant mediation effect in inferior frontal gyrus *pars opercularis* thickness for physical abuse and sexual abuse. This suggests that diverse forms of childhood trauma may largely converge on final common biological pathways involving smaller hippocampal and thinner prefrontal brain regions. This supports the idea that it is the cumulative stress burden or allostatic load, rather than the exact nature of the trauma, that drives its neurobiological consequences ^53^. In our sample, trauma subtype severities were correlated, so the effect of individual trauma types might be difficult to disentangle. Specific effects of single trauma dimensions are however suggested by the findings of fusiform gyrus and inferior frontal gyrus *pars opercularis* thickness mediating the effects of specific trauma types. The fusiform gyrus is key for emotional recognition, which is altered in neglected children and adolescents, and previous studies also found an association effect of childhood trauma on the fusiform gyrus^37,54^. It is intriguing that functional differences in this region have also been observed in individuals with bipolar disorder ^55^. Future studies should confirm whether this indirect pathway through the fusiform gyrus is specific to emotional trauma, potentially using emotion processing tasks that selectively engage this region.

Here, we focused on the association between childhood trauma and bipolar disorder. However, it is possible that altered gray matter induced by childhood trauma may be a transdiagnostic risk factor for psychopathology ^56^. Childhood trauma is a well-known risk factor for numerous psychiatric conditions, and hippocampal and prefrontal alterations have been documented across several diagnoses, such as major depression, schizophrenia and post-traumatic stress disorder ^57–62^, as well as in psychiatrically healthy people ^63^. Future research should investigate brain structure mediators of childhood trauma across diagnostic categories to determine whether the same brain alterations play a role in other conditions. It will also be important to assess whether these effects are truly specific to childhood trauma, or whether other early stressors (e.g., poverty, illness, family dynamics) produce similar neurobiological effects mediating an increased likelihood of a participant having bipolar disorder.

Our findings raise the intriguing possibility that interventions that prevent or restore gray matter loss in the hippocampus, medial orbitofrontal cortex and superior frontal gyrus may reduce the psychiatric burden of childhood trauma. For example, preclinical and clinical studies suggest that some pharmacological or neuromodulation treatments may stimulate hippocampal neurogenesis and even reverse stress-induced hippocampal shrinkage ^64^. Specifically in bipolar disorder, chronic lithium use is associated with larger hippocampal volume and neurogenesis ^65^. One could speculate that treatments such as lithium might disrupt the link between early trauma and later development of psychopathology, including bipolar disorder, or remediate structural and functional alterations. Focusing on the regions we identified in future mechanistic analyses could potentially facilitate the development of novel targeted therapies.

In sum, the strengths of our study are the largest sample to date, our novel HIMA approach, and the fact that our findings were robust to leave-one-site-out cross-validation across 19 sites. Still, our work has limitations. First, our cross-sectional design prevents us from drawing definitive conclusions about causality ^66^. While smaller hippocampal volume and thinner frontal cortex may be a consequence of childhood trauma that in turn elevates risk for bipolar disorder, establishing causality of such effects requires longitudinal studies tracking children from trauma exposure through to illness onset. Another consideration is the heterogeneity of our sample, which spanned multiple sites and included individuals with varied demographics, environment, illness symptomatology, age of onset, durations, medication exposures, and comorbidities of related psychiatric conditions. All these demographic and clinical factors could have potentially confounded our results, although we believe that our leave-one-site-out cross-validation approach and the convergence of our findings with prior studies, at least in part, mitigate this concern. Another limitation is the retrospective self-report of childhood trauma, which does not always agree with prospective assessment ^67^. Finally, part of the direct association between childhood trauma and bipolar disorder could be explained by mediators that we were not able to capture in this analysis, such as medication use ^68^, illness duration, illness severity, comorbidities, mood state or heterogeneous clinical features within bipolar disorder (e.g. anxious distress, melancholic features, rapid cycling, etc.). ENIGMA-BD is continuing to collect deeper phenotyping information and to standardize clinical measures such as mood state, illness severity and comorbidities, across sites. This will lead to the ability to build more precise models, which may capture additional mediators or improve measurement precision of the mediators we identified.

In conclusion, our results show that childhood trauma is associated with an increased risk of a bipolar disorder diagnosis, in part via its association with lower gray matter in the hippocampus, medial orbitofrontal gyrus and superior frontal gyrus. Therefore, these results identified a potential biological pathway that may link childhood trauma, brain structure and risk for developing bipolar disorder. This could pave the way for a better mechanistic understanding of the illness, facilitate the identification of those individuals at-risk of trauma-related psychopathology and enable the development of new interventions before onset or during the course of the illness.

## Data availability

The data underlying this work are only available to members of the ENIGMA consortium.

