## Supplementary Material for "Brain morphology mediators of the link between childhood trauma and bipolar disorder: a large-scale international analysis"

Leonardo Tozzi, MD, PhD¹; Maria R. Dauvermann, PhD²; Emma Corley, PhD³; Andrea Fernandes, MS³; Melody J. Y. Kang⁴; Yanghee Im MS⁴; Leila Nabulsi, PharmD, PhD³^,^⁴; Roberto Goya-Maldonado, MD⁵; Klaus Berger, MD,PhD⁶; Marco Hermesdorf, PhD⁶; Hilary P. Blumberg, MD⁷; Lejla Colic, PhD⁷^,^⁸^,^⁹; Tamsyn E. Van Rheenen, PhD¹⁰^,^¹¹; Elysha Ringin, PhD¹⁰; Susan L. Rossell, PhD¹¹; James A. Karantonis, PhD¹⁰; Lisa S. Furlong, MNeuroSci¹⁰; Tilo Kircher, MD¹²; Frederike Stein, PhD¹²; Udo Dannlowski, MD, PhD¹³^,^¹⁴; Dominik Grotegerd, PhD¹³; Jair C. Soares, MD, PhD¹⁵; Mon-Ju Wu, PhD¹⁵; Giovana B. Zunta-Soares, MD¹⁵; Benson Mwangi, PhD¹⁵; Joaquim Radua, MD, PhD¹⁶; Enric Vilajosana, MS¹⁶; Eduard Vieta MD, PhD¹⁶; Melissa J. Green, PhD¹⁷; Emilie Olie, MD, PhD¹⁸; Guillaume Clain, MS¹⁹; Francesco Benedetti, MD²⁰^,^²¹; Elisa Melloni, PhD²⁰; Beatrice Bravi, PhD²⁰; Delfina Janiri, MD²²^,^²³; Daniela Vecchio, PhD²⁴; Fabrizio Piras, PhD²⁴; Nerisa Banaj, PhD²⁴; Gabriele Sani, MD²²^,^²³; Gloria Roberts, PhD¹⁷; Janice M. Fullerton, PhD²⁵^,^²⁶; Bronwyn J. Overs²⁶; Philip B. Mitchell MBBS MD¹⁷; Jonathan Savitz²⁷, PhD; Lakshmi N. Yatham, MBBS²⁸; Josselin Houenou, MD, PhD¹⁰^,^²⁹; Marion Leboyer, MD, PhD³⁰; Amanda Rodrigue, PhD³¹; David C. Glahn, PhD³¹; Sophia I. Thomopoulos, BA⁴; Neda Jahanshad, PhD⁴; Paul M. Thompson, PhD⁴; Ole A. Andreassen, MD, PhD³²; Christopher R. K. Ching, PhD⁴; Dara M. Cannon PhD³; Yann Quidé, PhD³³^,^³⁴; and The ENIGMA Bipolar Disorder Working Group

Affiliations:

1. Department of Psychiatry and Behavioral Sciences, Stanford University School of Medicine, Stanford, USA
2. Institute for Mental Health, School of Psychology, University of Birmingham, Birmingham, United Kingdom
3. Clinical Neuroimaging Laboratory, Galway Neuroscience Center, College of Medicine, Nursing and Health Sciences, University of Galway, Galway, Ireland
4. Imaging Genetics Center, Mark and Mary Stevens Neuroimaging & Informatics Institute, Keck School of Medicine, University of Southern California, Marina del Rey, CA, USA
5. Laboratory of Systems Neuroscience and Imaging in Psychiatry (SNIP-Lab), Department of Psychiatry and Psychotherapy, University Medical Center Göttingen, Göttingen, Germany
6. Institute of Epidemiology and Social Medicine, University of Münster, Münster, Germany
7. Department of Psychiatry, Yale School of Medicine, New Haven, USA
8. Department of Psychiatry and Psychotherapy, Jena University Hospital, Jena, Germany
9. German Center for Mental Health (DZPG), Partner Site Halle-Jena-Magdeburg, Germany
10. Department of Psychiatry, Faculty of Medicine, Dentistry and Health Sciences, University of Melbourne, Melbourne, Australia
11. Center for Mental Health and Brain Sciences, Swinburne University, Melbourne, Australia
12. Department of Psychiatry and Psychotherapy, Marburg University, Marburg, Germany
13. Institute for Translational Psychiatry, University of Münster, Germany
14. Bielefeld University, Medical School and University Medical Center OWL, Protestant Hospital of the Bethel Foundation, Department of Psychiatry, Germany
15. Center of Excellence on Mood Disorders, Louis A. Faillace, MD, Department of Psychiatry and Behavioral Sciences, McGovern Medical School, University of Texas Health Science Center at Houston, TX, USA
16. IDIBAPS, Hospital Clinic Barcelona, CIBERSAM–Instituto de Salud Carlos III, Institute of Neuroscience, Universitat de Barcelona, Barcelona, Catalonia, Spain
17. School of Clinical Medicine, The University of New South Wales (UNSW) Sydney, Sydney, Australia
18. IGF, University of Montpellier, CNRS, INSERM, Montpellier, France
19. Institut d’Imagerie Fonctionnelle Humaine, Platform I2FH, Department of Neuroradiology, University Hospital of Montpellier, Montpellier, France
20. Psychiatry & Clinical Psychobiology, Division of Neuroscience, IRCCS Ospedale San Raffaele, Milan, Italy
21. University Vita-Salute San Raffaele, Milan, Italy
22. Department of Neuroscience, Section of Psychiatry, Università Cattolica del Sacro Cuore, Rome, Italy
23. Department of Neuroscience, Section of Psychiatry, Fondazione Policlinico Universitario Agostino Gemelli IRCCS, Rome, Italy
24. Neuropsychiatry Laboratory, Department of Clinical Neuroscience and Neurorehabilitation, Santa Lucia Foundation IRCCS, Rome, Italy
25. School of Biomedical Sciences, Faculty of Medicine & Health, University of New South Wales, Sydney, Australia
26. Neuroscience Research Australia, Randwick, Sydney, NSW, Australia
27. Laureate Institute for Brain Research, Tulsa, OK, USA
28. Department of Psychiatry, University of British Columbia, Vancouver, Canada
29. Mental Health Services, Northern Health, Epping, Australia
30. INSERM U955 Team "Translational Neuropsychiatry," Université Paris-Est Créteil, Créteil, France
31. Department of Psychiatry, Boston Children’s Hospital, Harvard Medical School, Boston, MA, USA
32. Center for Precision Psychiatry, University of Oslo, Oslo University Hospital, Oslo, Norway
33. NeuroRecovery Research Hub, School of Psychology, The University of New South Wales (UNSW) Sydney, Sydney, NSW, Australia
34. Center for Pain IMPACT, Neuroscience Research Australia, Randwick, NSW, Australia

Supplementary Material

### Supplementary Tables

**Supplementary Table 1: Inclusion/Exclusion Criteria**

| **Sample** | **Depression Measure** | **Instrument for diagnosis of BD/MDD** | **Inclusion/Exclusion criteria** |
| --- | --- | --- | --- |
| AFFDIS | MADRS, HDRS, BDI | Patients diagnosed by their psychiatrists with a depressive phase of major depressive disorder (UD group) or a depressive phase of bipolar disorder (BD group) (cf. ICD-10) and sex, age, and education matched HC were recruited by announcements at the local University Psychiatric Hospitals. | All participants were between 18 and 65 years old. Exclusion criteria were contraindications for MRI scans (i.e., pregnancy, metallic implants), strong visual impairment, neurological disorder, brain surgery or head trauma, current substance abuse, past or present psychiatric disorder for HC, and comorbid psychiatric diagnosis other than anxiety disorders for BD and UD. |
| BiDirect | HDRS, CES-D | M.I.N.I. Neuropsychiatric Interview, IDS, HAMD, CESD, ICD-10 | All participants were aged 35 to 65 years, participants in the depression cohort were recruited during inhospital treatment for a depressive episode, excluded were those with a concurrent substance use disorder, classification into BD type was done by the treating psychiatrist. |
| CHRM2 | HDRS | Structured Clinical Interview for DSM-IV | CHRM2 Study: Inclusion criteria were 18-65 years of age. A diagnosis of BD (and Euthymia) was confirmed using the Diagnostic and Statistical Manual of Mental Disorders (DSM-IV-TR) Structured Clinical Interview for DSM Disorders (American Psychiatric Association, 1994) conducted by an experienced psychiatrist. Exclusion criteria included neurological disorders, learning disability, comorbid misuse of substance/alcohol and of axis-1 disorders, history of head injury resulting in loss of consciousness for >5 minutes along with a history of oral steroid use in the previous 3 months. Mood symptoms severity was assessed using the Hamilton Anxiety (HARS) and Depression (HDRS-21) Rating Scale and the Young Mania Rating Scale (YMRS) at MRI scanning. Healthy controls had no personal history of a psychiatric illness or history among first-degree relatives, defined using the Structured Clinical Interview for DSM-IV – Non-patient edition (American Psychiatric Association, 1994). |
| COGSBD | MADRS | MINI-BD | All participants were aged 18-70. Exclusion criteria for all participants included; a history of neurological and neurodegenerative conditions, visual and hearing impairments, history of severe head injury, substance abuse/dependence in the past three months, and difficulties with written or spoken English. Furthermore, BD participants were excluded if they had non-trivial medication changes in the two months prior to assessment, or benzodiazepine use in the 48 hours prior. Healthy controls were excluded if they had a personal or family history of psychiatric illness, or current/previous psychotropic medication use. |
| FOR2107-Marburg | HDRS, BDI | Structured Clinical Interview for DSM-IV-TR for Axis I Diagnoses | Inclusion criteria: age 18-65 years; patients were diagnosed of bipolar I disorder by SCID-I Interview, currently depressed, (hypo)manic or remitted. Exclusion criteria all: any MRI contraindications; any neurological abnormalities. Exclusion criteria controls: any current or former psychiatric disorder; Exclusion criteria patients: substance dependence or current benzodiazepine treatment (wash out of at least three half-lives before study participation). |
| FOR2107-Muenster | HDRS, BDI | Structured Clinical Interview for DSM-IV-TR for Axis I Diagnoses | Inclusion criteria: age 18-65 years; patients were diagnosed of bipolar I disorder by SCID-I Interview, currently depressed, (hypo)manic or remitted. Exclusion criteria all: any MRI contraindications; any neurological abnormalities. Exclusion criteria controls: any current or former psychiatric disorder; Exclusion criteria patients: substance dependence or current benzodiazepine treatment (wash out of at least three half-lives before study participation). |
| Houston | MADRS, HDRS, BDI | Structured Clinical Interview for DSM-IV for Axis I Diagnoses | The study was approved by the Institutional Review Board of The University of Texas Health Science Center at San Antonio. Healthy controls reporting current or past Axis I disorders, suicidal history and a first-degree relative with any Axis I disorder were excluded. Participants with any endocrinological disease, head trauma, neurological disease, and family history of any hereditary neurological disorder or medical conditions such as hypertension, diabetes, active liver disease and kidney problems were excluded from this study. |
| IDIBAPS | HDRS | M.I.N.I. Neuropsychiatric Interview | The inclusion criteria require a diagnosis of bipolar disorder confirmed by two psychiatrists through a structured diagnostic interview. Exclusion criteria include: i) age younger than 18 or older than 65; ii) inability to provide consent; iii) participants with a history of head trauma or neurological disease; iv) individuals with a substance use disorder within 6 months prior to the scanning session (excluding nicotine). |
| IGP | MADRS | Diagnosis was confirmed using the OPCRIT algorithm applied to interviewer ratings on the DIP, acc. to ICD-10 criteria | All participants were fluent English speakers and aged 18-65 years old. In addition to general MR contra-indication, exclusion criteria included an inability to communicate sufficiently in English, a current neurological disorder, life- time head injury with loss of consciousness, a diagnosis of substance abuse or dependence in the past six months; and/or having been treated with electroconvulsive therapy in the previous six months. |
| MNC | MADRS, HDRS, BDI | Structured Clinical Interview for DSM-IV for Axis I Diagnoses | Inclusion criteria: age 17-65 years; patients were diagnosed of bipolar I disorder by SCID-Interview, currently depressed (HAMD >= 18); Exclusion criteria all: any MRI contraindications; any neurological abnormalities; Exclusion criteria controls: any current or former psychiatric disorder; Exclusion criteria patients: substance-related disorders or current benzodiazepine treatment (wash out of at least three half-lives before study participation), and former electroconvulsive therapy. |
| Montpellier | HDRS,BDI | M.I.N.I. Neuropsychiatric Interview | All participants were Caucasian right-handed (as assessed by the Edinburgh scale ) females. All participants had to be euthymic at the time of scanning, as indicated by a Hamilton Depression Rating Scale score <7 and a Young Mania Rating Scale score < 7. Other exclusion criteria were a lifetime history of severe head trauma, CNS disorder, schizophrenia, and a history of alcohol or drug abuse or dependence within the past 12 months.  Healthy controls had no past or present psychiatric disorder. |
| Olin | MADRS, HDRS | Structured Clinical Interview for DSM-IV for Axis I Diagnoses | Study 1- Patients were identified through outpatient clinics and community mental health facilities in the Hartford area. Inclusion criteria for patients were aged between 18 and 70 years, diagnosis of bipolar I disorder as determined by the Structured Clinical Interview for DSM-IV (SCID), and no first degree relatives with a bipolar disorder diagnosis. Unrelated healthy comparison subjects were included if they had no lifetime history of axis I psychiatric disorder as assessed by the SCID and no family history of mood or psychotic disorders. Participants were excluded for alcohol or drug abuse or dependence within the past 6 months, a history of major medical or neurological disorders, or IQ <70 as assessed by the WAIS. In patients, euthymia was established with the Hamilton Depression Rating Scale (HAM-D), the Young Mania Rating Scale, the Brief Psychiatric Rating Scale (BPRS), and through diagnostic case reviews.    Study 2- Inclusion criteria: ages 15-65 years; proficiency in English at the sixth-grade level or higher; no significant neurologic disorders including those secondary to head injury; no history of substance abuse within the last month or substance dependence within the last 6 months; and negative urine toxicology screening results on the day of testing. The healthy controls met the following additional criteria: no personal or family history (first degree) of psychotic or bipolar disorders; no personal history of recurrent mood disorder; no lifetime history of substance dependence; and no history of any significant cluster A Axis II personality features defined by meeting full criteria or within 1 criterion of a cluster A diagnosis using the Structured Interview for DSM-IV Personality. |
| OSR-Milano | HDRS, IDS | Structured Clinical Interview for DSM-IV FOR AXIS I DISORDERS | Exclusion criteria were age younger than 18; the presence of other diagnoses on Axis I; the presence of pregnancy; history of epilepsy or major medical, neurological disorders or brain trauma; history of drug or alcohol abuse or dependency. Patients were also required to have a current IQ in the normal range (>70). No patient had received electroconvulsive therapy within 6 months prior to study enrolment. |
| Rome | MADRS, HDRS, BDI | Structured Clinical Interview for DSM-IV-TR Axis I Disorders, Patient Edition (SCID-I/P) | Outpatients with DSM-IV-TR diagnoses of BD type I (BD-I) and BD type II (BD-II), were recruited at the Sant’Andrea clinic and IRCCS Santa Lucia Foundation outpatient clinics in Rome, Italy. inclusion criteria were as follows: (a) age 18-65 years; (b) at least 5 years of education; (c) able to undergo magnetic resonance imaging (MRI); (d) Italian language native speaker; (e) at least 6 months of stable pharmacotherapy for BD. Exclusion criteria were: (a) diagnosis of substance dependence or abuse in the 2 years before the assessment; (b) traumatic brain injury with loss of consciousness; (c) major medical or neurological conditions; (d) Mini-Mental State Examination (MMSE) score lower than 24 (given that scores below this level are suggestive of cognitive deterioration based on normative data from the Italian population); (e) left-handedness; and (f) any MRI-identified brain abnormality or microvascular lesion on T1-weighted, T2-weighted images, or fluid attenuated inversion recovery (FLAIR) sequences; (g) incomplete or incorrect scan segmentation.  HCs were recruited from the same geographical area. All HCs were screened for lifetime personal history of DSM-IV-TR Axis I and II disorders using the SCID-I/NP and SCID-II as well as for family history (up to 2nd degree relatives) of mood disorders or schizophrenia. Participants with DSM-IV-TR Axis I or II disorders and/or family history of mood disorders or schizophrenia were excluded from the HC group. All other eligibility criteria were the same as those for the BD group. |
| Sydney | MADRS | Diagnostic Interview for Genetic Studies (for 22-30 year-olds); Kiddie-SADS (for 12-21 year-olds) | Bipolar disorder participants meet DSM-IV criteria for either bipolar I or bipolar II disorder. Control participants meet inclusion criteria if no parent or sibling had bipolar I or II disorder, recurrent major depression, schizoaffective disorder, schizophrenia, recurrent substance abuse or any past psychiatric hospitalization; and no parent with a first degree relative had a past mood disorder hospitalization or history of psychosis. All subjects were aged between 12 and 30 years. For those aged between 12 and 21 an adapted version of the Schedule for Affective Disorders and Schizophrenia for School-Age Children – Present and Lifetime Version (K-SADS-BP) was developed specifically for use in the US-Australia collaborative study of young people at genetic risk for BD. For participants aged between 22 and 30 the DIGS (Version 4) is used to measure the current and lifetime presence of axis I DSM-IV disorders. |
| Tulsa | MADRS, HDRS, IDS | Structured Clinical Interview for DSM-IV | Bipolar disorder participants met DSM-IV criteria for either bipolar I or bipolar II disorder, or BD NOS. Age 18-55. The unmedicated BD group did not receive any psychotropic medications for at least 3 weeks (8 for fluoxetine) prior to the MRI scanning. The healthy control individuals met the same exclusion criteria except that they had no personal or family (first-degree relatives) history of psychiatric illness assessed using the Structured Clinical Interview for the DSM-IV-TR and the Family Interview for Genetic Studies (FIGS). Exclusion criteria were as follows: serious suicidal ideation or behavior; medical conditions or concomitant medications likely to influence CNS or immunological function including cardiovascular, respiratory, endocrine and neurological diseases; a history of drug or alcohol abuse within 6 months or a history of drug or alcohol dependence within 1 year (DSM-IV-TR criteria), and general MRI exclusion criteria such as magnetic implants or claustrophobia. |
| UBC | MADRS, HDRS | Mini-International Neuropsychiatric Interview (MINI) for DSM-IV and ICD-10 | Inclusion criteria were: aged 14 to 35 years; first episode mania within 3 months of enrollment. Inclusion criteria were deliberately broad to capture a wide range of clinical presentations. Exclusion criteria were: inability to take part in neuropsychological testing; meets the standard criteria for exclusion for magnetic resonance imaging; a previous manic episode diagnosed retrospectively on structured interview or via collateral. Age- and sex-matched healthy controls were also recruited, with the following exclusion criteria for healthy controls: personal history of psychiatric disorder; family history of any major psychiatric disorder in the first or second degree relatives; inability to take part in neuropsychological testing; meets the standard criteria for exclusion for magnetic resonance imaging. |
| VIP | NA | Diagnostic Interview for Genetic Studies | Inclusion criteria for study participation were ages between 18 and 65, no history of alcohol or drug abuse/dependence, no history of mental retardation, no previous head trauma with loss of consciousness, and no current or past cardiac or neurological disease. We excluded subjects with any significant cerebral anatomic anomaly. In addition, HC were free of any personal past or present personal psychiatric disorder and first-degree family history of schizophrenia, schizoaffective disorder or BD. Participants were not included for MRI if MRI was contraindicated or if pregnant. |
| Yale - Blumberg | HDRS | Structured Clinical Interview for DSM-IV for Axis I Diagnoses | Inclusion criteria for all participants were ages between 18 and 60 years. Inclusion criteria for the BD group were meeting diagnostic criteria for BD according to the Diagnostic and Statistical Manual of Mental Disorders (Fourth ed. Text Revision; DSM-IV-TR), while for the HC group inclusion criteria were absence of any Axis I psychiatric disorders or first-degree relative with a major mood or psychotic disorder, assessed with the Family History Screen for Epidemiological Studies (Lish et al., 1995). Exclusion criteria for all participants included MRI contraindications and medical and neurological illnesses and conditions (including loss of consciousness >5 min) that could alter cerebral tissue, except treated thyroid disorders in 5 BD participants. |

**Supplementary Table 2: Imaging acquisition**

| **Cohort** | **Scanner type** | **Sequence T1** | **FreeSurfer version** | **Slice orientation** |
| --- | --- | --- | --- | --- |
| AFFDIS | 3T Siemens Magnetom TrioTim | 3D T1 (176 slices; TR = 2250 ms; TE = 3.26 ms; FOV 256; voxel size 1X1X1mm) | 5,3 | Sagittal |
| BiDirect | 3T Philips Intera | 3D T1-weighted turbo field echo (TFE). TR=7.26 ms; TE=3.56 ms; flip angle=9º; voxel size= 1x1x1 | 5,3 | Sagittal |
| CHRM2 | 3T Philips Achieva | 3D T1-weighted MPRAGE. Voxel size= 0.83 x 0.83 x 0.89. Slices: 180. TE= 3.046, TR= 8.5. Flip angle= 8°. | 5,3 | Axial |
| COGSBD | 3T Siemens Magnetom TrioTim | 3D T1 weighted sequence. TR=1900 msec; TE=2.52 msec; flip angle=9°, 176 slices, voxel size 1x1x1 | 6 | Sagittal |
| FOR2107-Marburg | 3T Siemens Magnetom Trio | 3D T1-weighted magnetization prepared rapid acquisition on gradient echo (MPRAGE). TR= 2130ms, TE= 2.28, flip angle= 9 degrees. 176 slices. | 5,3 | Sagittal |
| FOR2107-Muenster | 3T Siemens PRISMA | 3D T1-weighted magnetization prepared rapid acquisition on gradient echo (MPRAGE). TR= 1900ms, TE= 2.26, flip angle= 8 degrees. 192 slices. | 5,3 | Sagittal |
| Houston | 1.5T Philips Gyroscan Intera | 3D T1-weighted spoiled gradient recalled acquisition in steady state. 144 slices. Voxel size = 1x1x1. Flip angle 40°. TR= 2, TE= 5. T1= 220ms. | 5,3 | Transverse |
| IDIBAPS | 3T Siemens Magnetom Prisma fit | 3D T1-weighted magnetization prepared rapid acquisition on gradient echo (MPRAGE). 240slices. Voxel size= 0.9*0.9*1. TI= 900ms.TE= 3.01ms, TR= 2300ms. Flip angle= 9º. | 5,3 | Sagittal |
| IGP | 3T Achieva Philips TX | 3D T1- weighted magnetization prepared rapid acquisition gradient echo (MPRAGE) (200 slices; TR = 8.9ms; TE = 4.1ms, voxel size 0.9x0.9x0.9 | 5,3 | Sagittal |
| MNC | 3T Philips Gyroscan Intera | 3D Fast gradient echo sequence. 320 slices. Voxel size= .5 x .5 x .5. TI= 815. TR= 7.4, TE= 3.4 Flip angle= 9° | 5,3 | Coronal |
| Montpellier | 1,5T Siemens Magneton Avento | 3D T1 (160 slices; TR = 2100 ms; TE = 4.1 ms; flip angle=15º; voxel size 1X1X1mm) | 6 | Axial |
| Olin | 3T Siemens Allegra | 3D T1-weighted magnetization prepared rapid acquisition gradient echo (MPRAGE). Flip angle=9°, repetition time=2.3 milliseconds, echo time=2.91 milliseconds; voxel size 1x1x1.2 mm; 160 slices. | 5,1 | Sagittal |
| OSR-Milano | 3T Philips Ingenia and 3T Philips Intera scanner | 3D-MPRAGE sequence: TR 2500 ms, TE 4.6 ms, field of view FOV = 230 mm, matrix = 256 × 256, in-plane resolution 0.9 × 0.9 mm, yielding 220 transversal slices with a thickness of 0.8 mm. | 5,3 | Axial |
| Rome | 3T Achieva MR imager (Philips) | Whole-brain T1-weighted images were obtained in the sagittal plane using a MPRAGE sequence (TE/TR = 5.4/11 ms, flip angle 8°, voxel size 0.54 × 0.54 × 0.9 mm3, FH(mm) 230; AP(mm) 233.594; RL(mm) 171, acquisition time 4 minutes and 26 seconds, using the SENSE acceleration | 5,3 | Sagittal |
| Sydney | 3T Philips Achieva | 3D T1-weighted turbo field echo (TFE). Slices 180. Voxel size 1x1x1. TR= 5.5, TE= 2.5. Flip angle= 8° | 5,3 | Sagittal |
| Tulsa | 3T GE MR750 Discovery | 3D T1-weighted magnetization prepared rapid acquisition gradient echo (MPRAGE). Flip angle=8°, repetition time=6 milliseconds, echo time=2.01 milliseconds; voxel size .938x0.938x0.9mm; 186 slices. | 5,3 | Axial |
| UBC | Philips Achieva 3.0 Tesla scanner | Whole-brain T1-weighted magnetic resonance images. FOV = 20 cm (RL) × 25.6 cm (AP), ACQ matrix = 256 × 256, isotropic image voxels (1 × 1 × 1 mm3) TR/TE = autoset shortest, T/R head coil, flip angle = 8  degrees, and 1 mm thick contiguous 180 slices of the whole brain | 7,1 | Axial |
| VIP | 3T Siemens Tim Trio | 3D T1-weighted magnetization prepared rapid acquisition gradient echo (MPRAGE). Flip angle=9°, repetition time=2.3 milliseconds, echo time=2.98 milliseconds; voxel size 1x1x1.1 mm; 160 slices. | 5,3 | Sagittal |
| Yale-Blumberg | 3 T Siemens Trio | A high resolution, three-dimensional Magnetization Prepared Rapid Acquisition Gradient Echo (MPRAGE) T1-weighted sequence with parameters: repetition time=1500 ms, echo time=2.77 ms, matrix=256 × 256, field of view=256 × 256mm2, 160 1 mm slices without gap for a voxel-size of 1mm3, and two averages. | 6 | Sagittal |

**Supplementary Table 3: Missing values in the dataset as a result of missing questionnaires and exclusion of gray matter measures after quality control.** We show the number and % of participants missing each variable used in the analysis.

| **Variable** | **Count** | **Proportion** |
| --- | --- | --- |
| R entorhinal thickness | 391 | 12% |
| R entorhinal surface | 387 | 12% |
| L superiortemporal surface | 383 | 12% |
| L bankssts thickness | 382 | 12% |
| L superiortemporal thickness | 382 | 12% |
| L bankssts surface | 382 | 12% |
| L middletemporal thickness | 305 | 9% |
| L middletemporal surface | 303 | 9% |
| L entorhinal thickness | 296 | 9% |
| R superiortemporal thickness | 290 | 9% |
| R superiortemporal surface | 287 | 9% |
| L entorhinal surface | 283 | 9% |
| L cuneus surface | 277 | 9% |
| R medialorbitofrontal thickness | 271 | 8% |
| L medialorbitofrontal thickness | 264 | 8% |
| R medialorbitofrontal surface | 260 | 8% |
| L cuneus thickness | 259 | 8% |
| L inferiortemporal thickness | 252 | 8% |
| L medialorbitofrontal surface | 248 | 8% |
| R supramarginal thickness | 246 | 8% |
| L temporalpole thickness | 245 | 8% |
| L supramarginal thickness | 241 | 7% |
| R pericalcarine thickness | 241 | 7% |
| L pericalcarine surface | 241 | 7% |
| L inferiortemporal surface | 240 | 7% |
| R supramarginal surface | 233 | 7% |
| L pericalcarine thickness | 232 | 7% |
| R bankssts thickness | 231 | 7% |
| L supramarginal surface | 231 | 7% |
| R pericalcarine surface | 229 | 7% |
| L temporalpole surface | 227 | 7% |
| R bankssts surface | 227 | 7% |
| R cuneus thickness | 225 | 7% |
| L pallidum volume | 219 | 7% |
| R temporalpole thickness | 218 | 7% |
| R cuneus surface | 218 | 7% |
| R insula thickness | 208 | 6% |
| R temporalpole surface | 202 | 6% |
| R insula surface | 201 | 6% |
| R inferiortemporal thickness | 195 | 6% |
| L lingual thickness | 194 | 6% |
| R inferiorparietal thickness | 194 | 6% |
| L lingual surface | 194 | 6% |
| R lingual thickness | 190 | 6% |
| L rostralanteriorcingulate thickness | 188 | 6% |
| R middletemporal thickness | 185 | 6% |
| R inferiortemporal surface | 184 | 6% |
| R postcentral thickness | 183 | 6% |
| R inferiorparietal surface | 182 | 6% |
| L postcentral thickness | 180 | 6% |
| L insula thickness | 179 | 6% |
| R lateralorbitofrontal thickness | 179 | 6% |
| L rostralanteriorcingulate surface | 177 | 5% |
| R lingual surface | 177 | 5% |
| R middletemporal surface | 177 | 5% |
| R rostralanteriorcingulate thickness | 173 | 5% |
| R precentral thickness | 172 | 5% |
| L lateralorbitofrontal thickness | 171 | 5% |
| L inferiorparietal thickness | 170 | 5% |
| R postcentral surface | 170 | 5% |
| R rostralanteriorcingulate surface | 170 | 5% |
| R lateraloccipital thickness | 169 | 5% |
| L postcentral surface | 166 | 5% |
| L insula surface | 165 | 5% |
| R lateralorbitofrontal surface | 163 | 5% |
| L lateraloccipital thickness | 162 | 5% |
| L precentral thickness | 158 | 5% |
| L inferiorparietal surface | 158 | 5% |
| R lateraloccipital surface | 158 | 5% |
| R precentral surface | 158 | 5% |
| L superiorfrontal thickness | 155 | 5% |
| L lateralorbitofrontal surface | 155 | 5% |
| L caudalanteriorcingulate thickness | 153 | 5% |
| R superiorparietal thickness | 150 | 5% |
| L parstriangularis thickness | 148 | 5% |
| L rostralmiddlefrontal thickness | 147 | 5% |
| L superiorparietal thickness | 147 | 5% |
| L lateraloccipital surface | 147 | 5% |
| R parstriangularis thickness | 143 | 4% |
| L precentral surface | 143 | 4% |
| R isthmuscingulate thickness | 142 | 4% |
| R precuneus thickness | 142 | 4% |
| R rostralmiddlefrontal thickness | 141 | 4% |
| R superiorfrontal thickness | 140 | 4% |
| L fusiform thickness | 138 | 4% |
| R caudalanteriorcingulate thickness | 138 | 4% |
| R parsopercularis thickness | 138 | 4% |
| L superiorfrontal surface | 138 | 4% |
| R superiorparietal surface | 137 | 4% |
| L caudalanteriorcingulate surface | 136 | 4% |
| L caudalmiddlefrontal thickness | 135 | 4% |
| R isthmuscingulate surface | 135 | 4% |
| L parsorbitalis thickness | 134 | 4% |
| R frontalpole thickness | 134 | 4% |
| L frontalpole thickness | 133 | 4% |
| L parstriangularis surface | 133 | 4% |
| L rostralmiddlefrontal surface | 133 | 4% |
| L precuneus thickness | 132 | 4% |
| R posteriorcingulate thickness | 132 | 4% |
| L paracentral thickness | 131 | 4% |
| R precuneus surface | 131 | 4% |
| L superiorparietal surface | 130 | 4% |
| R parstriangularis surface | 130 | 4% |
| R parahippocampal thickness | 129 | 4% |
| R parsopercularis surface | 129 | 4% |
| L parahippocampal thickness | 128 | 4% |
| R paracentral thickness | 128 | 4% |
| R rostralmiddlefrontal surface | 128 | 4% |
| R superiorfrontal surface | 128 | 4% |
| L isthmuscingulate thickness | 127 | 4% |
| L parsopercularis thickness | 127 | 4% |
| R fusiform thickness | 127 | 4% |
| L paracentral surface | 127 | 4% |
| R caudalmiddlefrontal thickness | 126 | 4% |
| L posteriorcingulate thickness | 125 | 4% |
| L precuneus surface | 124 | 4% |
| R parahippocampal surface | 124 | 4% |
| R caudalanteriorcingulate surface | 123 | 4% |
| R parsorbitalis thickness | 122 | 4% |
| L fusiform surface | 122 | 4% |
| L parahippocampal surface | 122 | 4% |
| L transversetemporal thickness | 121 | 4% |
| R transversetemporal thickness | 120 | 4% |
| L caudalmiddlefrontal surface | 118 | 4% |
| L parsorbitalis surface | 118 | 4% |
| R frontalpole surface | 118 | 4% |
| L hippocampus volume | 116 | 4% |
| R paracentral surface | 116 | 4% |
| R posteriorcingulate surface | 116 | 4% |
| L frontalpole surface | 115 | 4% |
| L isthmuscingulate surface | 114 | 4% |
| L parsopercularis surface | 114 | 4% |
| L accumbens volume | 112 | 3% |
| R fusiform surface | 112 | 3% |
| R caudalmiddlefrontal surface | 111 | 3% |
| L posteriorcingulate surface | 109 | 3% |
| L transversetemporal surface | 109 | 3% |
| R parsorbitalis surface | 109 | 3% |
| R transversetemporal surface | 109 | 3% |
| R thalamus volume | 100 | 3% |
| R accumbens volume | 89 | 3% |
| L putamen volume | 83 | 3% |
| CTQ total | 78 | 2% |
| R caudate volume | 77 | 2% |
| L amygdala volume | 77 | 2% |
| R pallidum volume | 74 | 2% |
| R hippocampus volume | 72 | 2% |
| R putamen volume | 71 | 2% |
| R amygdala volume | 70 | 2% |
| L caudate volume | 68 | 2% |
| CTQ emotional neglect | 64 | 2% |
| CTQ physical neglect | 63 | 2% |
| CTQ emotional abuse | 61 | 2% |
| CTQ physical abuse | 61 | 2% |
| CTQ sexual abuse | 60 | 2% |
| L thalamus volume | 45 | 1% |
| Intracranial volume | 5 | 0% |
| Diagnosis | 0 | 0% |
| Age | 0 | 0% |
| Sex | 0 | 0% |
| Site | 0 | 0% |
| Site | 0 | 0% |

**Supplementary Table 4: Direct and indirect high-dimensional mediation model effects.** We show the mean, standard deviation (SD) and 95% confidence interval of the mean (CI) of each effect across 19 leave-one-site-out cross-validation folds retained in at least one fold. We also show in how many folds the effect was retained in the feature selection procedure. We considered significant effects for which the 95% CI of the mean across folds did not encompass 0. Age, sex and intracranial volume were also included as covariates in the mediation paths but are not shown here.

| Effect | Coefficient median (min-max) | 95% CI | Folds retained | p |
| --- | --- | --- | --- | --- |
| Direct effects |  |  |  |  |
| Childhood trauma | 0.841 (0.776, 0.893) | [0.834, 0.851] | 19 (100%) | <0.001 |
| Age | -0.037 (-0.164, 0.118) | [-0.046, -0.009] | 19 (100%) | <0.001 |
| Sex (male) | -0.192 (-0.273, -0.088) | [-0.202, -0.180] | 19 (100%) | <0.001 |
| Indirect effects |  |  |  |  |
| Hippocampus volume | 0.004 (0, 0.008) | [0.002, 0.005] | 15 (79%) | <0.001 |
| Medial orbitofrontal gyrus thickness | 0.002 (0, 0.004) | [0.002, 0.003] | 18 (95%) | 0.001 |
| Superior frontal gyrus thickness | 0.002 (0, 0.005) | [0.002, 0.003] | 11 (58%) | <0.001 |
| Fusiform gyrus thickness | 0 (0, 0.008) | [0, 0] | 6 (32%) | 1 |
| Inferior frontal gyrus *pars opercularis* thickness | 0 (0, 0.009) | [0, 0.003] | 6 (32%) | 1 |
| Inferior temporal gyrus thickness | 0 (0, 0.003) | [0, 0] | 5 (26%) | 1 |
| Lingual gyrus thickness | 0 (0, 0.001) | [0, 0] | 2 (11%) | 1 |
| Caudal middle frontal gyrus thickness | 0 (0, 0.001) | [0, 0] | 1 (5%) | 1 |
| Lingual gyrus surface | 0 (0, 0.001) | [0, 0] | 2 (11%) | 1 |
| Supramarginal gyrus surface | 0 (-0.001, 0) | [0, 0] | 1 (5%) | 1 |
| Caudate volume | 0 (-0.002, 0) | [0, 0] | 5 (26%) | 1 |

**Supplementary Table 5: Correlation between childhood trauma subtypes.** We show Spearman correlations between each pair of subscales of the Childhood Trauma Questionnaire (short form).

|  | Emotional abuse | Physical abuse | Sexual abuse | Emotional neglect | Physical neglect |
| --- | --- | --- | --- | --- | --- |
| Emotional abuse | 1.00 | 0.53 | 0.41 | 0.56 | 0.43 |
| Physical abuse | 0.53 | 1.00 | 0.38 | 0.38 | 0.35 |
| Sexual abuse | 0.41 | 0.38 | 1.00 | 0.29 | 0.28 |
| Emotional neglect | 0.56 | 0.38 | 0.29 | 1.00 | 0.57 |
| Physical neglect | 0.43 | 0.35 | 0.28 | 0.57 | 1.00 |

**Supplementary Table 6: Direct and indirect high-dimensional effects of mediation model using emotional neglect as predictor.** We show the mean, standard deviation (SD) and 95% confidence interval of the mean (CI) of each effect across 19 leave-one-site-out cross-validation folds retained in at least one-fold. We also show in how many folds the effect was retained in the feature selection procedure. We considered significant effects for which the 95% CI of the mean across folds did not encompass 0. Age, sex and intracranial volume were also included as covariates in the mediation paths but are not shown here.

| Effect | Coefficient median (min-max) | 95% CI | Folds retained | p |
| --- | --- | --- | --- | --- |
| Direct effects |  |  |  |  |
| Emotional neglect | 0.594 (0.561, 0.619) | [0.582, 0.601] | 19 (100%) | <0.001 |
| Age | -0.035 (-0.190, 0.016) | [-0.063, -0.017] | 19 (100%) | <0.001 |
| Sex | -0.106 (-0.188, -0.025) | [-0.134, -0.072] | 19 (100%) | <0.001 |
| Indirect effects |  |  |  |  |
| Hippocampus volume | 0.002 (0, 0.005) | [0.002, 0.004] | 16 (84%) | 0.001 |
| Medial orbitofrontal gyrus thickness | 0.002 (0, 0.004) | [0.001, 0.002] | 16 (84%) | <0.001 |
| Superior frontal gyrus thickness | 0.003 (0, 0.006) | [0, 0.003] | 13 (68%) | <0.001 |
| Fusiform gyrus thickness | <0.001 (0, 0.006) | [0, 0.003] | 10 (53%) | 0.001 |

**Supplementary Table 7: Direct and indirect high-dimensional effects of mediation model using emotional abuse as predictor.** We show the mean, standard deviation (SD) and 95% confidence interval of the mean (CI) of each effect across 19 leave-one-site-out cross-validation folds retained in at least one fold. We also show in how many folds the effect was retained in the feature selection procedure. We considered significant effects for which the 95% CI of the mean across folds did not encompass 0. Age, sex and intracranial volume were also included as covariates in the mediation paths but are not shown here.

| Effect | Coefficient median (min-max) | 95% CI | Folds retained | p |
| --- | --- | --- | --- | --- |
| Direct effects |  |  |  |  |
| Emotional abuse | 0.804 (0.754, 0.840) | [0.793, 0.810] | 19 (100%) | <0.001 |
| Age | 0.075 (-0.021, 0.117) | [0.045, 0.090] | 19 (100%) | <0.001 |
| Sex | -0.288 (-0.367, -0.208) | [-0.319, -0.260] | 19 (100%) | <0.001 |
| Indirect effects |  |  |  |  |
| Hippocampus volume | 0.003 (0, 0.008) | [0.002, 0.004] | 18 (95%) | <0.001 |
| Medial orbitofrontal gyrus thickness | 0.003 (0, 0.007) | [0.003, 0.004] | 19 (95%) | <0.001 |
| Superior frontal gyrus thickness | 0.004 (0, 0.012) | [0, 0.005] | 13 (68%) | <0.001 |
| Fusiform gyrus thickness | 0.001 (0, 0.008) | [0, 0.003] | 11 (58%) | 0.001 |

**Supplementary Table 8: Direct and indirect high-dimensional effects of mediation model using physical neglect as predictor.** We show the mean, standard deviation (SD) and 95% confidence interval of the mean (CI) of each effect across 19 leave-one-site-out cross-validation folds retained in at least one fold. We also show in how many folds the effect was retained in the feature selection procedure. We considered significant effects for which the 95% CI of the mean across folds did not encompass 0. Age, sex and intracranial volume were also included as covariates in the mediation paths but are not shown here.

| Effect | Coefficient median (min-max) | 95% CI | Folds retained | p |
| --- | --- | --- | --- | --- |
| Direct effects |  |  |  |  |
| Physical neglect | 0.461 (0.410, 0.488) | [0.451, 0.466] | 19 (100%) | <0.001 |
| Age | -0.031 (-0.160, 0.034) | [-0.043, -0.010] | 19 (100%) | <0.001 |
| Sex | -0.045 (-0.155, 0.024) | [-0.102, -0.033] | 19 (100%) | <0.001 |
| Indirect effects |  |  |  |  |
| Hippocampus volume | 0.002 (0.001, 0.003) | [0.001, 0.002] | 19 (100%) | <0.001 |
| Medial orbitofrontal gyrus thickness | 0.001 (0, 0.002) | [0.001, 0.001] | 18 (95%) | <0.001 |
| Superior frontal gyrus thickness | 0.001 (0, 0.004) | [0, 0.002] | 14 (74%) | <0.001 |
| Fusiform gyrus thickness | 0.001 (0, 0.005) | [0, 0.002] | 12 (63%) | <0.001 |

**Supplementary Table 9: Direct and indirect high-dimensional effects of mediation model using physical abuse as predictor.** We show the mean, standard deviation (SD) and 95% confidence interval of the mean (CI) of each effect across 19 leave-one-site-out cross-validation folds retained in at least one fold. We also show in how many folds the effect was retained in the feature selection procedure. We considered significant effects for which the 95% CI of the mean across folds did not encompass 0. Age, sex and intracranial volume were also included as covariates in the mediation paths but are not shown here.

| Effect | Coefficient median (min-max) | 95% CI | Folds retained | p |
| --- | --- | --- | --- | --- |
| Direct effects |  |  |  |  |
| Physical abuse | 0.437 (0.365, 0.490) | [0.431, 0.446] | 19 (100%) | <0.001 |
| Age | 0.003 (-0.105, 0.125) | [-0.010, 0.026] | 19 (100%) | <0.001 |
| Sex | -0.084 (-0.163, 0.047) | [-0.114, -0.063] | 19 (100%) | <0.001 |
| Indirect effects |  |  |  |  |
| Hippocampus volume | 0.003 (0, 0.009) | [0.002, 0.004] | 15 (79%) | <0.001 |
| Medial orbitofrontal gyrus thickness | 0.002 (0, 0.004) | [0.001, 0.002] | 17 (89%) | <0.001 |
| Inferior frontal gyrus pars opercularis thickness | 0.002 (0, 0.007) | [0.001, 0.003] | 16 (84%) | <0.001 |

**Supplementary Table 10: Direct and indirect high-dimensional effects of mediation model using sexual abuse as predictor.** We show the mean, standard deviation (SD) and 95% confidence interval of the mean (CI) of each effect across 19 leave-one-site-out cross-validation folds retained in at least one fold. We also show in how many folds the effect was retained in the feature selection procedure. We considered significant effects for which the 95% CI of the mean across folds did not encompass 0. Age, sex and intracranial volume were also included as covariates in the mediation paths but are not shown here.

| Effect | Coefficient median (min-max) | 95% CI | Folds retained | p |
| --- | --- | --- | --- | --- |
| Direct effects |  |  |  |  |
| Sexual abuse | 0.587 (0.505, 0.677) | [0.581, 0.593] | 19 (100%) | <0.001 |
| Age | 0.011 (-0.114, 0.073) | [-0.011, 0.030] | 19 (100%) | <0.001 |
| Sex | -0.220 (-0.317, -0.143) | [-0.263, -0.193] | 19 (100%) | <0.001 |
| Indirect effects |  |  |  |  |
| Hippocampus | 0.003 (0, 0.007) | [0, 0.004] | 11 (58%) | <0.001 |
| Inferior frontal gyrus pars opercularis thickness | <0.001 (-0.002, 0.001) | [-0.001, 0] | 19 (100%) | <0.001 |

**Supplementary Table 11: Direct and indirect high-dimensional mediation model effects when excluding participants containing any missing values.** We show the median, minimum, maximum and 95% confidence interval of the median of each significant effect across 19 leave-one-site-out cross-validation folds. We chose the median rather than the mean to account for the zero-inflated values generated by regularization. We also show in how many folds the effect was retained in the feature selection procedure. To determine significance, we generated a null distribution of the effects by permutation testing, used it to calculate a two-sided p-value and considered significant effects with p<0.05.

| Effect | Median (min, max) | 95% CI | Folds retained | p |
| --- | --- | --- | --- | --- |
| Direct effects |  |  |  |  |
| Childhood trauma | 0.841 (0.776, 0.893) | [0.834, 0.851] | 19 (100%) | <0.001 |
| Age | -0.037 (-0.164, 0.118) | [-0.046, -0.009] | 19 (100%) | <0.001 |
| Sex (male) | -0.192 (-0.273, -0.088) | [-0.202, -0.180] | 19 (100%) | <0.001 |
| Indirect effects |  |  |  |  |
| Hippocampus volume | 0.004 (0, 0.008) | [0.002, 0.005] | 15 (79%) | <0.001 |
| Medial orbitofrontal gyrus thickness | 0.002 (0, 0.004) | [0.002, 0.003] | 18 (95%) | <0.001 |
| Superior frontal gyrus thickness | 0.002 (0, 0.008) | [0, 0.005] | 11 (58%) | <0.001 |

**Supplementary Table 12: Direct and indirect high-dimensional mediation model effects when excluding participants under 18 years of age.** We show the median, minimum, maximum and 95% confidence interval of the median of each significant effect across 19 leave-one-site-out cross-validation folds. We chose the median rather than the mean to account for the zero-inflated values generated by regularization. We also show in how many folds the effect was retained in the feature selection procedure. To determine significance, we generated a null distribution of the effects by permutation testing, used it to calculate a two-sided p-value and considered significant effects with p<0.05.

| Effect | Median (min, max) | 95% CI | Folds retained | p |
| --- | --- | --- | --- | --- |
| Direct effects |  |  |  |  |
| Childhood trauma | 0.850 (0.782, 0.903) | [0.841, 0.863] | 18 (100%) | <0.001 |
| Age | 0.022 (-0.125, 0.167) | [0.001, 0.028] | 18 (100%) | <0.001 |
| Sex (male) | -0.167 (-0.277, -0.061) | [-0.188, -0.136] | 18 (100%) | <0.001 |
| Indirect effects |  |  |  |  |
| Hippocampus volume | 0.002 (0, 0.007) | [0, 0.005] | 12 (67%) | 0.001 |
| Medial orbitofrontal gyrus thickness | 0.002 (0, 0.005) | [0.002, 0.003] | 17 (94%) | <0.001 |
| Superior frontal gyrus thickness | 0.003 (0, 0.008) | [0, 0.005] | 11 (61%) | <0.001 |

### Supplementary Figures

**Supplementary Figure 1: Location of participating sites.**

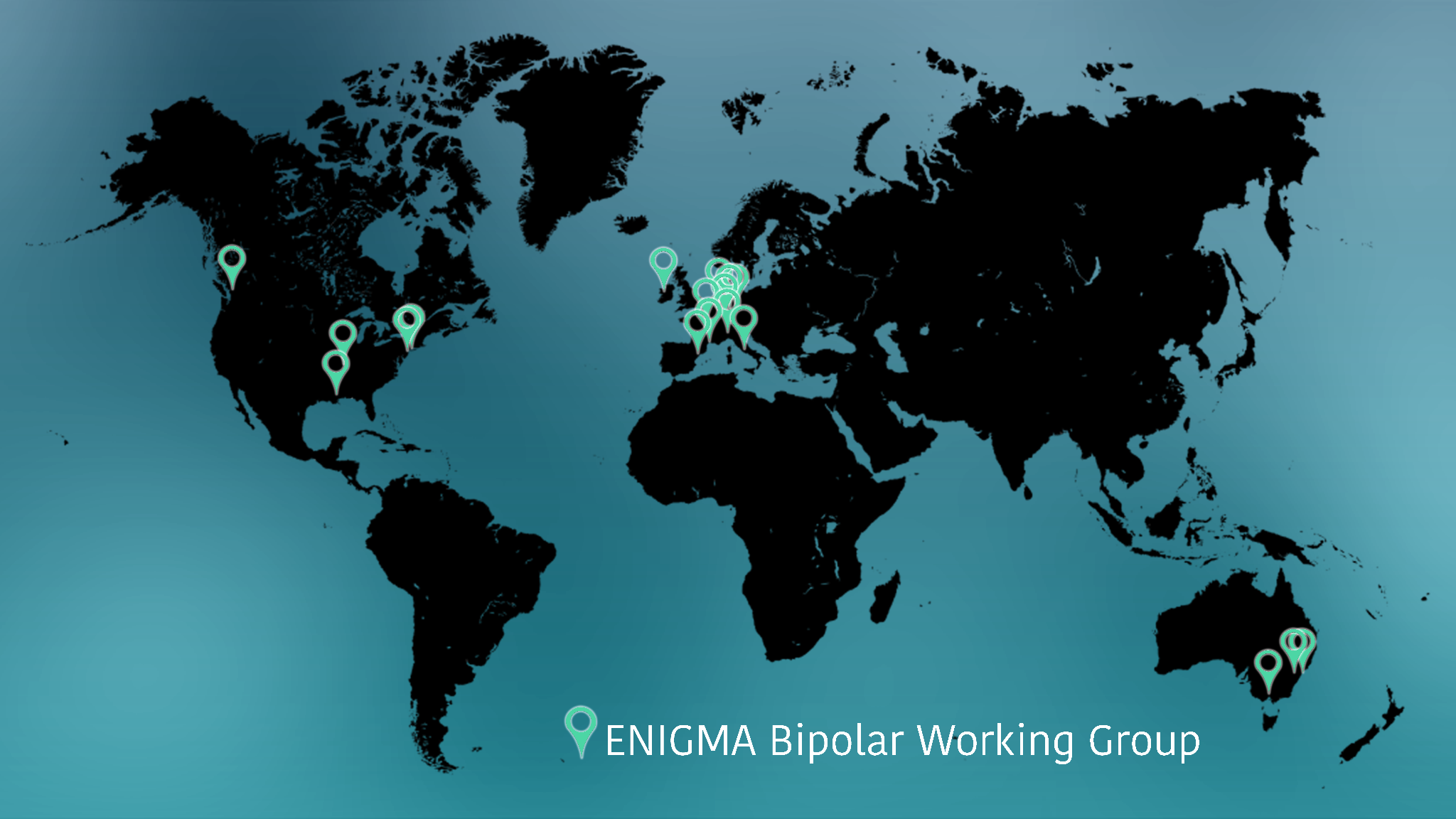

**Supplementary Figure 2: Area under the curve of the model for predicting bipolar disorder at each site.** We show as a red dashed line the mean AUC (0.71) and the confidence interval of the mean AUC as a shaded red area (0.67-0.76). AUC=area under the curve.

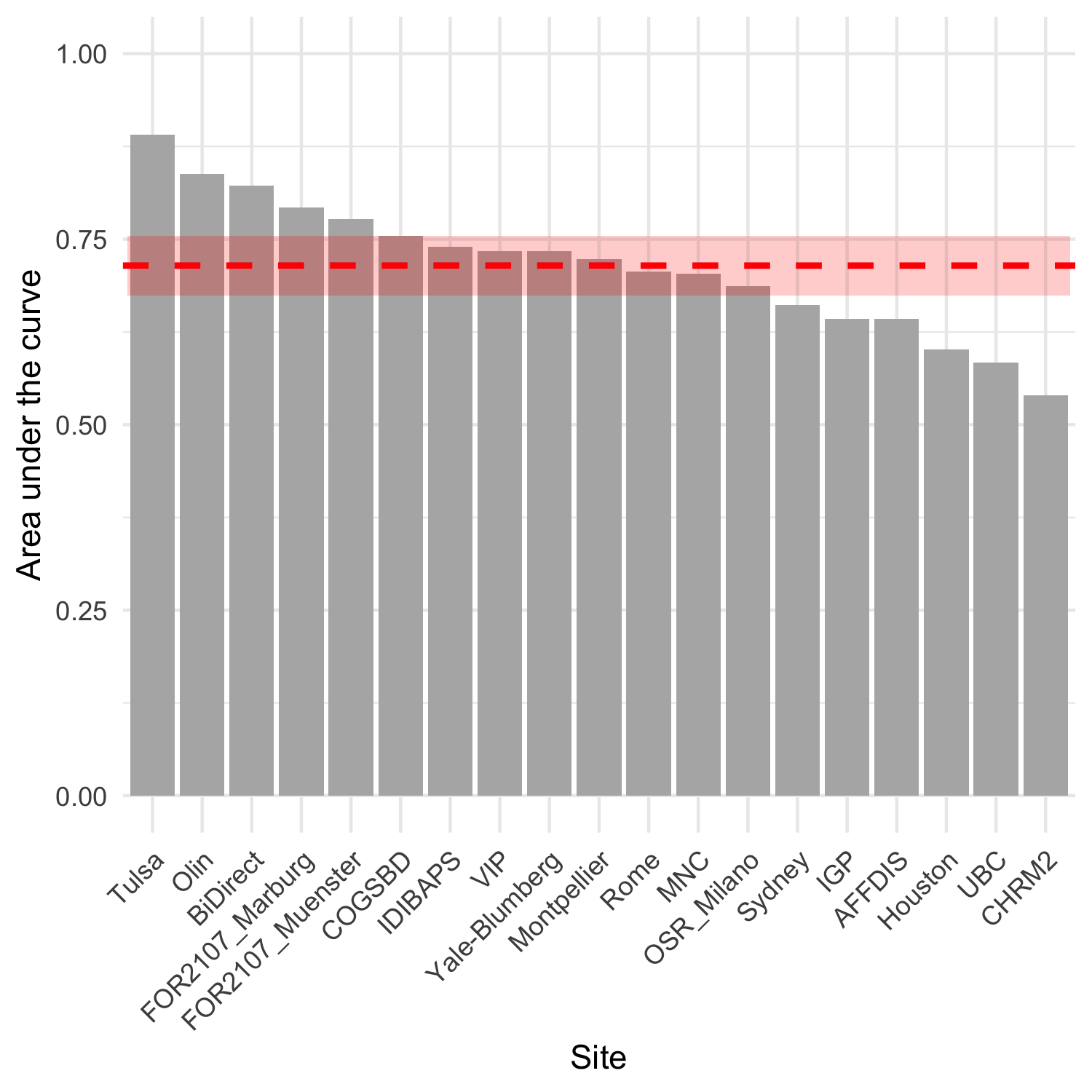
